# Genome-wide survey of parent-of-origin specific associations across clinical traits derived from electronic health records

**DOI:** 10.1101/2020.12.08.20246199

**Authors:** Hye In Kim, Bin Ye, Jeffrey Staples, Anthony Marcketta, Chuan Gao, Regeneron Genetics Center, Geisinger Regeneron DiscovEHR Collaboration, Alan R. Shuldiner, Cristopher V. Van Hout

**Author notes:** Correspondence: Hye In Kim, Cristopher V. Van Hout.

## Abstract

Parent-of-origin (PoO) effects refer to the differential phenotypic impact of genetic variants dependent on their parental inheritance. Genetic variants in imprinted genes can have PoO specific effects on complex traits, but these effects may be poorly captured by models that do not differentiate the parental origin of the variant. The aim of this study was to screen genome-wide imputed sequence for PoO effects on electronic health records (EHR) derived clinical traits in 134,049 individuals of European ancestry from the DiscovEHR study.

Using pairwise kinship estimates from genetic data and demographic data, we identified 22,051 offspring with at least one parent present in the DiscovEHR study. We then assigned the PoO of ∼9 million variants in the heterozygous offspring using two methods. First, when one of the parental genotypes was homozygous, we determined PoO based on apparent Mendelian segregation. Second, we estimated PoO by comparing parental and offspring haplotypes around the variant allele. Using these PoO assignments, we performed genome-wide PoO association analyses across 154 quantitative traits including lab test results and biometric measures and 612 binary traits of ICD10 3-digit codes extracted from EHR in the DiscovEHR study. Out of 732 PoO associations meeting a significance threshold of P <5×10^−8^, we attempted to replicate 274 PoO associations in the UK Biobank study, consisting of 462,453 individuals and including 5,015 offspring with at least one parent, and replicated 9 PoO associations with nominal significance threshold P <0.05.

In summary, the current study characterizes PoO effects of genetic variants genome-wide on a broad range of clinical traits derived from EHR in a large population study enriched for familial relationships. Our results suggest that 1) PoO specific effects are frequently captured by a standard additive model and that 2) statistical power to detect PoO specific effects remains modest even in large studies. Nonetheless, accurately modeling PoO effects of genetic variants has the potential to improve our understanding of the mechanism of the association and finding new associations that are not captured by the additive model.

## Introduction

Genomic imprinting, the non-equal expression of the two parental alleles, can lead to parent-of-origin (PoO) specific effects of genetic variants on traits. For example, when a gene is silenced on the paternally inherited chromosome and only expressed from the maternally inherited chromosome, functional variants of the gene that are maternally inherited may have observable phenotypic impacts while those that are paternally inherited may not, leading to maternal-specific effects. Imprinting has been characterized in approximately 1% of the genome and can influence complex traits in PoO specific manner^1,2^ and contribute to their heritability.

There have been studies that aimed to find PoO effects of genetic variants on traits. Many studies focused on associations identified using standard additive models in or near imprinted genes and then examined whether the additive signals are capturing PoO effects^2-7^. Other studies tested for PoO effects genome-wide to identify novel signals for specific traits, including type 2 diabetes (T2D), height, and autism spectrum disorder^2,8,9^. Recently, a study performed genome-wide PoO association analyses for 21 common quantitative traits in a Hutterite population isolate^10^. These studies have confirmed the presence of PoO effects in multiple loci including imprinted *KCNQ1, KLF14, IGF2, DLK1*, and *GNAS* loci^2-8,11^, and observed diverse patterns of PoO effects including simple uniparental effects and differential effects between the parental alleles.

Extending previous efforts, we performed a genome-wide screen for PoO effects over a broad spectrum of clinical traits in the DiscovEHR study consisting of 134,049 individuals of European ancestry. The DiscovEHR study is enriched for parent-offspring relationships, allowing the assignment of PoO in as many as 22,051 individuals. This study systematically screens for PoO associations across the genome and across hundreds of EHR derived traits and can provide the baseline for future studies of PoO specific effects.

## Results

### Identification of offspring in DiscovEHR study

To find parent-offspring relationships among the 134,049 European ancestry DiscovEHR participants, we estimated the genome-wide identity-by-decent (IBD) in all pairs of individuals using array genotype data. There were 28,562 pairs with genome-wide probability of sharing one allele IBD (IBD1) greater than 0.8, which were inferred as parent-offspring relationships (Figure 1A). Then, we used age and sex information to assign father, mother, and offspring in these relationships. As a result, we found a total of 22,051 offspring, of which 4,896 had both parents, 11,815 had mothers, and 5,340 had fathers present in the study (Figure 1B).

**Figure 1.**
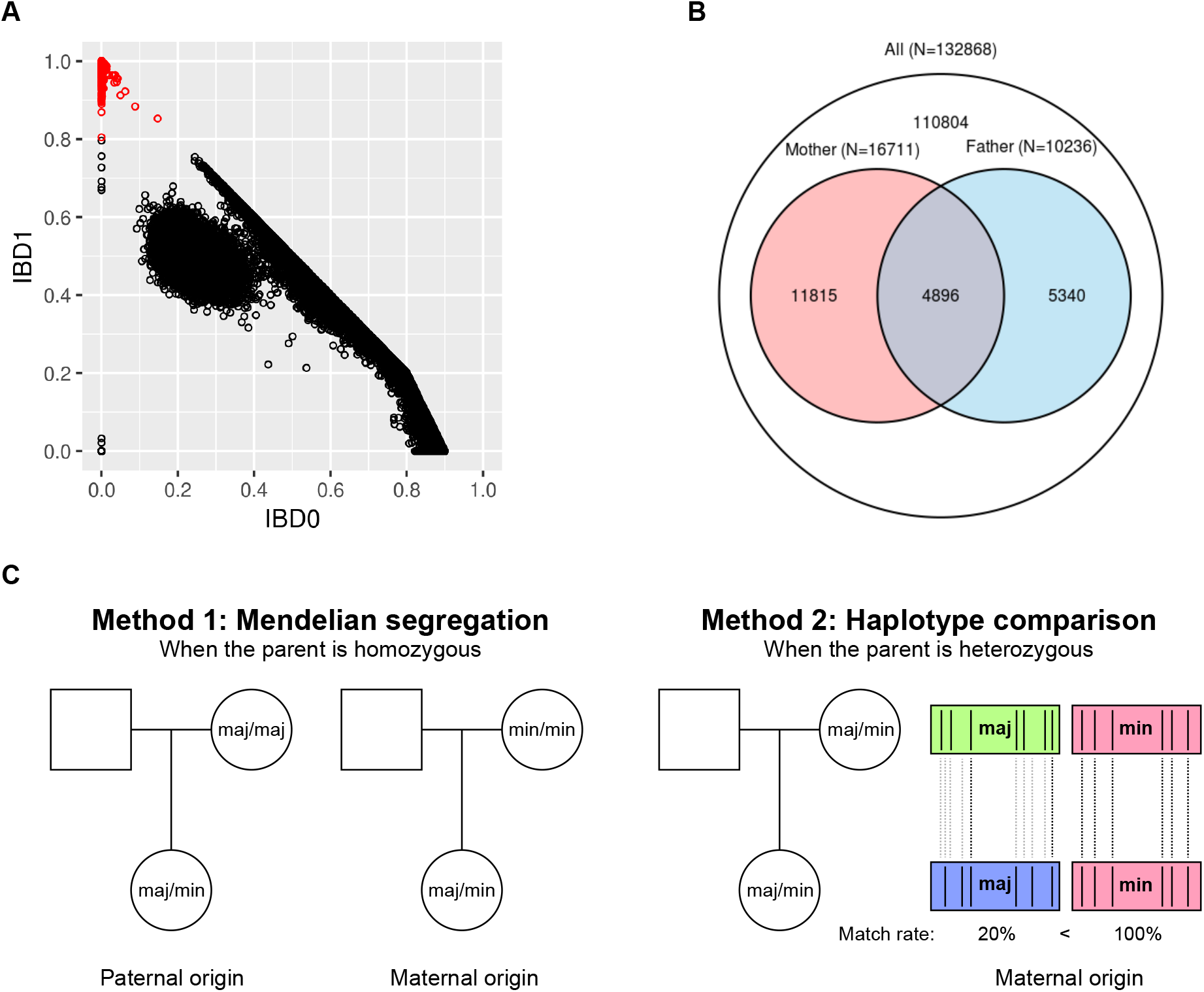
Identification of parent-offspring relationships and PoO assignment in DiscovEHR study. (A) Genome-wide identity-by-decent (IBD) was estimated from genetic data between every pair of individuals in the DiscovEHR study. Pairs with genome-wide probability of sharing one allele IBD (IBD1) >0.8 (in red) were inferred to be in parent-offspring relationships. (B) Based on age and sex information, father, mother, and offspring were inferred in each relationship. The number of offspring with one parent or both parents included in the study is indicated in the corresponding area of the Venn Diagram. (C) Among the heterozygous offspring of a given variant, parent-of-origin (PoO) of the minor allele was assigned using two methods. When the genotype of at least one parent was homozygous, PoO was determined based on Mendelian segregation. When the parental genotype(s) is/are heterozygous, PoO was estimated by comparing parental and offspring haplotypes (see text for detailed methods).

### Parent-of-origin (PoO) assignment of genetic variants in offspring

We assigned the PoO of a total of 9,085,657 imputed variants that passed quality control and had minor allele count (MAC) ≥10 among offspring and were LD-pruned (r^2^ <0.2). At each variant site, we assigned the PoO of the minor allele in the heterozygous offspring using two methods (Figure 1C). 1. Mendelian method: When the genotype of at least one parent was homozygous for either major or minor allele, PoO was determined based on Mendelian segregation. For example, if the genotype of the mother was homozygous major allele, then the PoO of the minor allele was inferred as paternal, while if the genotype of the mother was homozygous minor allele, then the PoO of the minor allele was inferred as maternal. 2. Haplotype method: Since the Mendelian method is uninformative when the parental genotype(s) is/are heterozygous, we also estimated PoO by comparing offspring and parental haplotypes within a 1Mb window that carry the same allele (major or minor) of the variant. The percent match rate, or percent of identical by state polymorphic nucleotides was calculated as below.

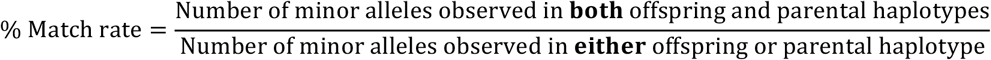

The allele that is carried on the offspring-parent haplotype pair that has a greater match rate was inferred as the allele that is inherited from the parent. We assessed the accuracy of the haplotype method for each variant by first applying it to the heterozygous offspring for whom PoO could be determined by the Mendelian method. As a quality control measure, when the PoO estimated by the haplotype method was <20% concordant with the PoO determined by the Mendelian method, only PoO determined by the Mendelian method were included in follow up analysis. Using this approach, we assigned the PoO of 9,085,586 and 8,801,949 variants (corresponding to 4,178,033,204 and 2,958,665,691 heterozygous genotypes) by the Mendelian and haplotype methods, respectively, leading to the overall PoO assignment rate of 99.2% of all heterozygous genotypes among the offspring. Among the 8,801,949 variants for which the haplotype method was used to estimate PoO, the overall concordance between the haplotype and Mendelian methods was 98.5%.

### Statistical models to identify PoO specific associations

To characterize PoO specific effects, we tested associations under parental and differential models as described below. Parental models include paternal and maternal models that contrast trait values in carriers of the paternally and maternally inherited allele, respectively, with homozygous individuals of the major allele. Heterozygous individuals with opposite or unknown PoO and homozygous individuals of the minor alleles are not included in parental models. Paternal- and maternal-specific associations were defined as being associated under one parental model and not the other. We also evaluated a differential model, which tests for a difference in trait values between paternally and maternally inherited alleles; contrasting heterozygous individuals of inferred paternal and maternal inheritance. While the parental models may offer greater power to detect PoO effects due to the number of homozygous major allele individuals, only the differential model directly measures differences between paternal and maternal allele inheritance.

### Testing PoO specificity of additive associations within imprinted regions

Functional variants that affect imprinted genes with monoallelic expression may be most likely to exert PoO specific effects on traits. Therefore, we examined whether variants with additive associations within the known imprinted regions have detectable PoO specific associations. We first tested 137,304 variants within a ±500kb window from 69 known or suggested imprinted genes^2,12^ (Supplementary Table 1) under the additive model across 173 quantitative traits derived from EHR (Supplementary Table 2) using BOLT linear mixed model^13^. These regions constitute approximately 1.3% of the genome. Six traits failed to produce results, due to the absence of estimated heritability. Among the remaining 167 traits, we found 667 additive associations that were statistically significant, P < 3.6×10^−7^, after Bonferonni correction for 137,304 variants. We tested these additive associations under paternal, maternal, and differential models and found that 12 had significant PoO specific associations, P < 7.5×10^−5^, after correction for 667 variant-trait associations: 4 were paternal-specific and 8 were maternal-specific associations, among which 4 also had differential associations (Table 1). Several of these associations were in LD with previously known PoO associations, serving as positive controls. For example, 7:130738173:T:C variant with maternal-specific association with HDL-C levels, TC/HDL-C ratios, and triglyceride levels is near the maternally expressed *KLF14* gene. The variant is in LD (r^2^=0.97) with rs4731702 that was previously found to have maternal-specific associations with *KLF14* gene expression and type 2 diabetes risk in Icelandic and American Indian populations^2,5^. While additive associations of *KLF14* locus with HDL-C and triglyceride levels have been reported, our results suggest a previously uncharacterized maternal-specific pattern. Another example is the maternal-specific association of 14:100704203:T:C variant with platelet counts near the paternally expressed *DLK1* gene, which replicates a previously reported association of rs7141210 (r^2^=0.81) in the Icelandic population^11^. This study showed that rs7141210 is associated with maternal-specific DNA methylation pattern, suggesting that the maternally inherited allele may impair the silencing of maternal gene expression. A third example is the maternal-specific association of 20:58872268:G:A variant with thyroid-stimulating hormone (TSH) levels near the maternally expressed *GNAS* gene, which confirms a previously reported association of rs139242164 (r^2^=0.62) in the Icelandic population^11^. Genetic variations in the *GNAS* gene are a well-established cause of pseudo-hypoparathyroidism (PHP) that is characterized by end-organ resistance to parathyroid hormone and high levels of circulating TSH^14^.

**Table 1.**
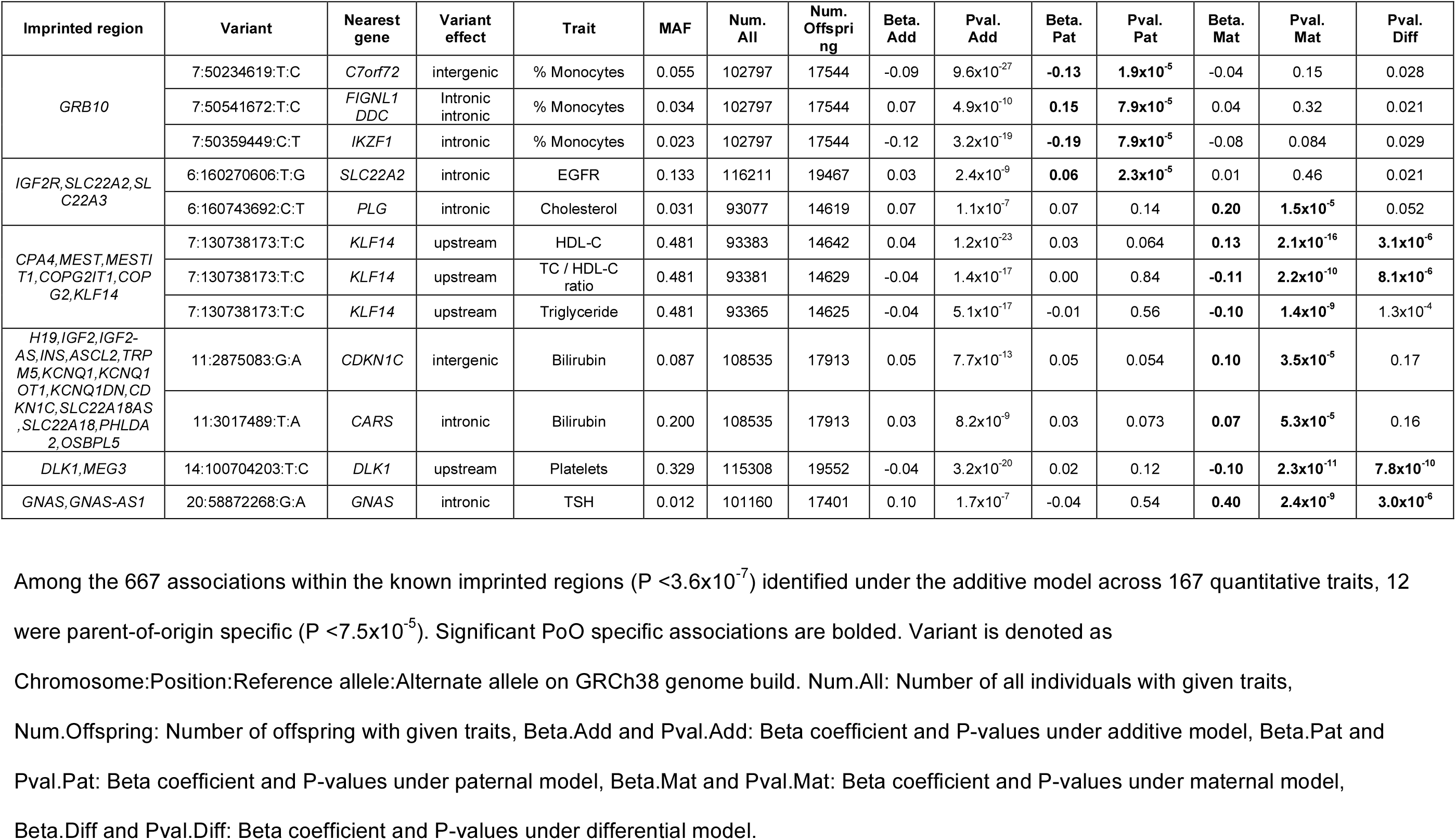
Parent-of-origin specificity among the additive associations identified within imprinted regions.

### Genome-wide PoO association analyses for 154 quantitative traits

To identify PoO specific effects outside the known imprinted regions, we performed genome-wide PoO association analyses for quantitative traits under parental and differential models using BOLT linear mixed model^13^. Among the 167 traits with non-zero estimated heritability, 13 traits with high genomic inflation (>1.5) under PoO models were omitted, yielding results for 154 traits. We found a total of 732 PoO associations (P <5×10^−8^) for 725 unique variants, including 341 paternal-specific, 344 maternal-specific, and 49 differential associations (Supplementary Tables 3-5). Notably, only 19 of these associations were within known imprinted regions. We attempted to replicate these associations in the UK Biobank study consisting of 462,453 individuals of European ancestry including 5,015 offspring with at least one parent. Of the 725 variants and 83 traits with associations in the DiscovEHR study, 721 variants and 37 traits could be unambiguously mapped in the UK Biobank study, allowing the examination of 274 associations for replication. We replicated 9 PoO associations (P <0.05): 4 were paternal-specific and 5 were maternal-specific associations, among which one also had differential association (Table 2). The strongest PoO association found was the aforementioned association of the variant near *DLK1* with platelet counts. Of the 9 PoO associations, 4 had stronger associations under the corresponding additive model, while the other 4 were only significant under the parental or differential model (P-values under additive model > 0.05).

**Table 2:**
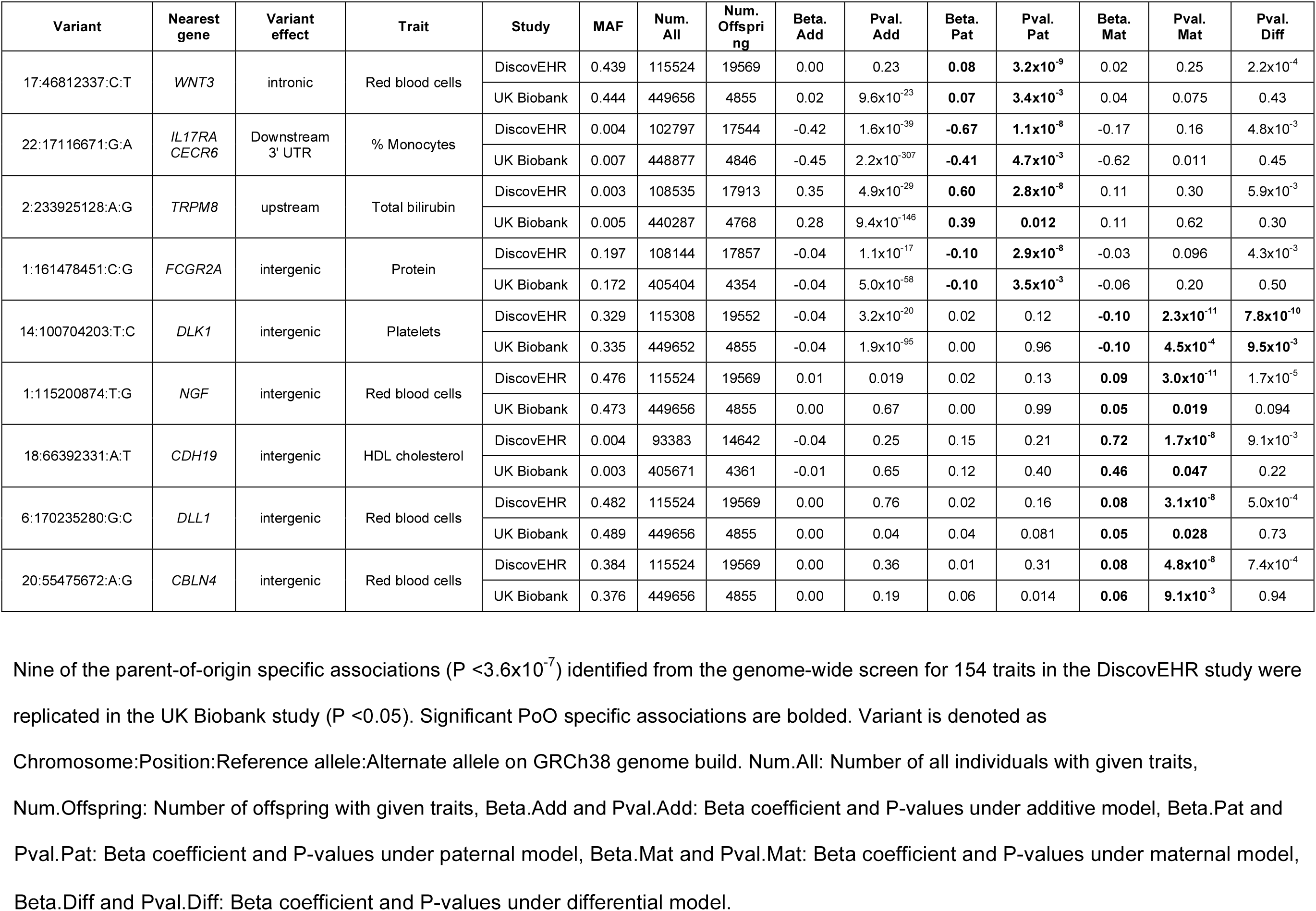
Parent-of-origin specific associations for quantitative traits identified in the DiscovEHR study (P <5×10^−8^) that are replicated in the UK Biobank study (P <0.05)

### Genome-wide PoO association analyses for 612 binary traits

We also conducted genome-wide PoO association analyses for binary traits based on ICD10 3-digit codes using SAIGE linear mixed model^15^. To mitigate computational burden required for testing association for binary traits, we tested the differential model as a screen for PoO effects genome-wide, and subsequently examined significant differential associations under additive and parental models. Of the 980 traits with at least 20 cases and 20 controls among the offspring, 612 traits were estimated to have non-zero heritability. All traits had genomic inflation factors below 1.5. We found 27 significant differential associations (P <5×10^−8^) (Supplementary Table 6), including two variants near the imprinted *IGF2* locus that were associated with type 2 diabetes (T2D). One of the associated variants, 11:1680825:A:T, is in LD (r^2^=0.84) with rs2334499 that was previously identified for PoO specific association with type 2 diabetes in the Icelandic population^2^. Consistent with the previous report, the paternal allele was associated with increased risk (OR=1.13, P=0.01) while the maternal allele was associated with decreased risk (OR=0.73, P=4.0×10^−10^), with significant differential association (P=8.5×10^−10^). The other variant, 11:1690902:G:A, was in low LD measured by r^2^, 0.17, with 11:1680825:A:T variant, but was in high D’ (0.98), indicating linkage disequilibrium. Interestingly, while it was the maternal allele of 11:1680825:A:T that was associated with reduced risk, the paternal allele of 11:1690902:G:A was associated with reduced risk. When attempting to replicate the 27 differential DiscovEHR associations for binary traits in the UK Biobank study, none reached nominal statistical significance (P <0.05), likely due to limited power based on low counts of cases and offspring in the UK Biobank study.

### Power simulations for PoO effects

Imprinting can give rise to diverse patterns of PoO effects of genetic variants: uniparental, polar dominance, and bipolar dominance^1^. We simulated the power to detect different types of PoO effects across ranges of effect sizes and minor allele frequencies (MAFs) with fixed parameters matching the DiscovEHR PoO analysis; study size and alpha (type 1 error). When a variant affects an imprinted gene with monoallelic expression from one parental allele, it can have uniparental (either maternal or paternal) effect on traits. Phenotypic impacts are observed in the heterozygous and homozygous offspring that inherited the variant from a specific parent, but not in the heterozygous individuals that inherited it from the other parent. (Figure 2A). In this case, the additive model has the greatest power, even though it does not take into account the phenotypic difference between heterozygous individuals with paternal and maternal inheritance. All heterozygous and homozygous individuals are included in the additive test, while parental and differential models only use heterozygous offspring with PoO assignment and exclude homozygous individuals. This is consistent with our observation that most of the PoO associations near the known imprinted genes with monoallelic expression had more significant P-values under the additive model than under PoO models (Table 1). Alternatively, when a variant exerts phenotypic impacts only in the heterozygous individuals that inherited the variant from a specific parent and not in the homozygous individuals or heterozygous individuals that inherited it from the other parent, the variant has a polar dominance effect (Figure 2B). Simulated polar dominance effects have the greatest power in additive or parental models, depending on the MAF of the variant. Specifically, the power of the additive model to capture polar dominance effects increases as the MAF increases up to ∼0.2, but beyond ∼0.2, the power diminishes because the larger number of homozygous individuals with no phenotypic alteration reduces effect estimates under the additive model. Instead, the parental model has greater power than the additive model when MAF is greater than ∼0.2. This may explain some of the paternal- and maternal-specific associations in Table 2 and Supplementary Tables 3 and 4 that have stronger associations under parental models than additive model. For example, a common intergenic variant near *NGF* (1:115200874:T:G, MAF=0.48) was significantly associated with red blood cell counts under the maternal model (beta=0.09, P=3.0×10^−11^), but only had weak association under the additive model (beta=0.01, P=0.019) (Table 2). Lastly, a bipolar dominance effect occurs when a variant has diverging phenotypic impacts between individuals of paternal and maternal inheritance without any impacts on homozygous individuals (Figure 2C). For this effect pattern, the differential model has the greatest power, even though it only utilizes heterozygous offspring with PoO assignment, because the effect estimates are largest when contrasting heterozygous offspring with paternal and maternal inheritance. In this case, the additive model has very limited power even though it utilizes the largest sample size because the effects of paternal and maternal allele will cancel each other and there are no phenotypic impacts on homozygous individuals. This may explain some of the differential associations in Supplementary Tables 5 and 6 that have stronger associations under differential model than under additive or parental models. For example, the association of a 3’ UTR variant of *ST8SIA5* (18:46674932:C:A) with total cholesterol to HDL cholesterol ratio was strongest under the differential model (P=1.4×10^−9^) and weaker under the parental models with opposite effects (beta=0.85, P=1.7×10^−6^ under paternal model and beta=-0.60, P=8.8×10^−5^ under the maternal model), but had no association under the additive model (beta=0.02, P=0.71) (Table S5).

**Figure 2.**
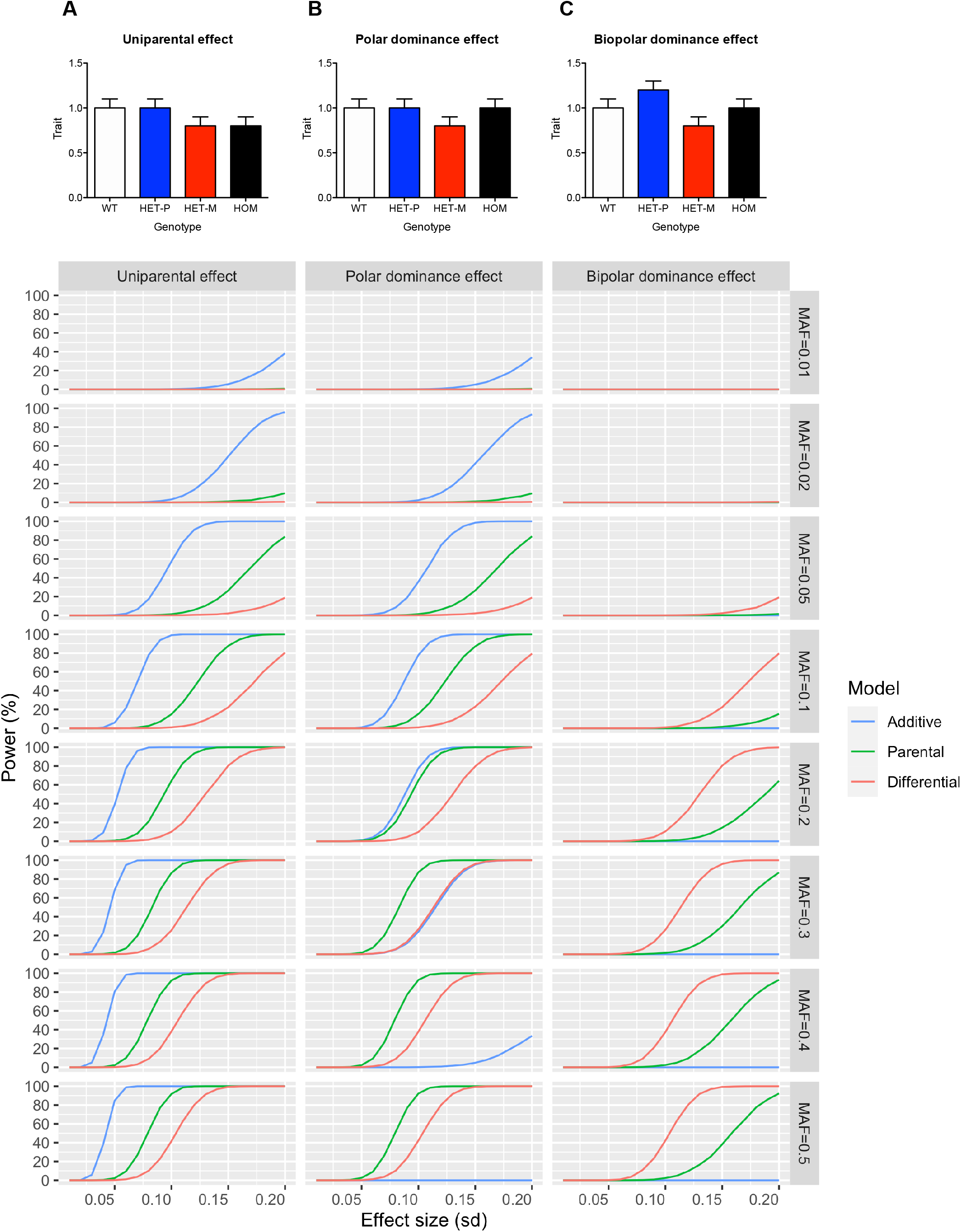
Simulation of power to detect PoO effects under different statistical models. Power to detect PoO effects under additive, parental, and differential models was simulated across ranges of minor allele frequencies (MAFs) and effect sizes assuming diverse patterns of PoO effects that could result from imprinting: (A) uniparental, (B) polar dominance, and (C) bipolar dominance effect. Bar plots at the top are illustrative examples of the various patterns of PoO effects that can result from imprinting. The horizontal axis displays a range of simulated effect sizes. The left vertical axes display the % power to detect the effect across a range of minor allele frequencies ordered on the right vertical. See text for detailed methods.

## Discussion

Variants that affect imprinted genes can have parent-of-origin (PoO) effects on traits and contribute to their variance; however, these effects may not be well captured by standard additive models that do not model parental origin. To address this, studies have employed approaches to assign parental origin of genetic variants based on known pedigrees and employed statistical models to specifically test for PoO effects, leading to the discovery of PoO specific associations for various traits^2,8-10^. The current study extends previous efforts by assigning the parental origin of genetic variants based on parent-offspring relationships inferred from genetic and demographic data in the absence of known pedigree and employing a high-throughput approach to detect PoO effects in a large, clinically ascertained study across hundreds of traits extracted from EHR.

From the genome-wide screen for PoO effects in the DiscovEHR study, we identified 732 PoO associations across 154 quantitative traits and 27 PoO associations across 612 binary traits. Many of the associations that we found in the DiscovEHR study could not be replicated in the UK Biobank study primarily due to the relatively modest number of offspring (5,015), as well as lack of matching traits for quantitative traits and limited number of cases for binary traits. This is evident by the observation that even well-established PoO associations with strong P-values in the DiscovEHR study, i.e. *KLF14* locus for lipids^2^, *GNAS* locus for TSH^11^ and *IGF2* locus for type 2 diabetes^2^ (Supplementary Tables 3-6), did not reach nominal significance for replication (P <0.05) in the UK Biobank study. Therefore, we anticipate that some of the associations that failed to replicate in the UK Biobank study may be replicated in future studies that are better powered. Power for detecting PoO effects can be enhanced by employing populations enriched for familial relationships and approaches that enable PoO assignment in a larger proportion of individuals. Additionally, approaches to leverage second-degree relatives for PoO assignment, such as in a study of the Icelandic population with extended pedigree information^2^, may further improve power. This may be possible in the absence of known pedigree structures by identifying second-degree relationships based on kinship estimates from genetic data, determining ancestors and descendants based on age, and assigning the parental side of the ancestors based on additional information such as mitochondrial DNA or Y chromosome.

Interestingly, of 9 associations that we discovered and replicated, only one association at the *DLK1* locus resides within the known imprinted regions. This raises the possibility that PoO effects may be more common and widespread than were previously thought. While around 100 genes in humans are known to be imprinted based on the currently available evidence^12,16^, high-throughput approaches applied to multiple tissue types in various developmental stages may reveal tissue- and time-specific imprinting effects on a larger number of genes. In line with this, recent studies that examined genome-wide PoO specific DNA methylation pattern suggested that DNA methylation pattern associated with imprinting is widespread and extends beyond the known imprinted regions^11,17^. In addition to imprinting, parental genetic effects where the genotypes of the parents directly affect the phenotype of the offspring, can lead to apparent PoO effects^18-20^. As we gain more evidence that PoO effects influence multiple complex traits, it would be important to estimate the extent to which PoO effects explain the variance of those traits.

Imprinting can give rise to diverse patterns of PoO effects of genetic variants on traits^1^, which in turn influence the power to detect these effects under different statistical models. Based on our power simulations, additive model has better power to detect associations resulting from uniparental effects than parental model. Nonetheless, parental models can provide more accurate effect size estimates for the causative parental allele, because the additive model would underestimate them by not differentiating the causative and non-causative parental alleles. In addition, parental and differential models can be better powered for detecting associations resulting from polar or bipolar dominance effects than additive model. This suggests that accurately accounting for the PoO effects of genetic variants can increase the statistical power and reserve the potential to identify novel association signals that are not captured under the additive model. Furthermore, knowing the PoO specific nature of genetic associations and effects can help accurately assess the risk conferred by the variants in the carriers and understand the molecular mechanism behind the genetic associations.

Many of the strongest PoO associations that we identified from the genome- and phenotype-wide scan for PoO effects in the DiscovEHR study show stronger additive associations and are suspected to result from uniparental effects (based on their proximity to known imprinted genes with monoallelic expression). This observation could suggest that additive model alone may be enough to capture most of the PoO effects with considerable contribution to genetic heritability and that it makes strategic sense to only test for additive association. While this may be true, we find it difficult to draw this conclusion empirically based on our current study given the limitation in the power and the small number of true positives. In addition, we did find many PoO associations with stronger associations under PoO models than under additive model, which may have resulted from either polar or bipolar dominance effects. We anticipate that future studies with greater statistical power and better mechanistic understanding of the PoO associations could help address this issue with more confidence.

In summary, we report a bioinformatic and statistical approach to screen for parent-of-origin (PoO) effects of genetic variants and its application in the DiscovEHR study enriched for familial relationships and phenotypic data derived from electronic health records. The current study provides a valuable reference point for future studies aimed to find parent-of-origin effects and suggests the need for approaches that can increase statistical power to detect PoO effects, methods to assess the contribution of PoO effects to genetic heritability of complex traits, and efforts to delineate the mechanisms behind the observed PoO associations by incorporating epigenetic and transcriptomic resources and experimental models.

## Methods

### Study participants and genetic data

The DiscovEHR study is a collaborative project between the Regeneron Genetics Center (RGC) and the Geisinger health system with participants enrolled in Geisinger’s MyCode Community Health Initiative^21^. All participants consented to provide genetic data and clinical data from their electronic health records for broad research use, including genetic analyses. The study was approved by the Institutional Review Board (IRB) at Geisinger. DNA samples from 143,575 individuals were genotyped at the RGC using either OmniExpress or Global Screening Array (GSA). 1,601 samples were excluded based on quality control criteria including gender mismatch, low call rate, discordancy compared to exomes, or suspected duplication. The array genotypes were imputed on the Michigan imputation server using Haplotype Reference Consortium (HRC) data as reference^22^. hg19 coordinates were converted to hg38 coordinates using Picard LiftoverVCF. Only individuals of European ancestry (N=134,049) as estimated by a previously described method^21^ and variants with imputation score ≥0.3 were used for subsequent analysis.

The UK Biobank study is a prospective cohort study consisting of around 500,000 individuals from the United Kingdom^23^. All participants consented the use of their genetic and medical information for research purpose. The study was approved by the North West Centre for Research Ethics Committee. DNA samples were genotyped using UK BiLEVE and UK Biobank Axiom arrays. The array genotypes were pre-phased by SHAPEIT3 and imputed by IMPUTE4 using HRC panel as reference^23^. As publicly available imputed sequence of the UK Biobank study did not retain phase information, the array genotypes of offspring and parents were imputed on the Michigan imputation server to obtain phased haplotypes which were used for PoO assignment as described in the following sections.

### Kinship estimation and identification of offspring

Genome-wide IBD proportions were estimated in all pairs of individuals using PLINK v1.9^24^ and array genotype data filtered by quality control metrics (individual missingness <0.1, variant missingness <0.1, Hardy-Weinberg equilibrium P <1×10^−15^), MAF (≥0.05), and LD-pruning (r^2^ <0.2). Pairs of individuals that have genome-wide IBD1 proportions >0.8 were inferred as parent-offspring relationships^25^. Age and sex information was used to identify offspring, father, and mother in these relationships.

### Parent-of-origin assignment of genetic variants in offspring

Imputed variants were filtered based on the quality control criteria (individual missingness <0.1, variant missingness <0.1, Hardy-Weinberg equilibrium P <1×10^−15^), minor allele count threshold (≥10 among the offspring), and LD-pruning (r^2^ <0.2) using PLINK2, resulting in 9,085,657 variants for PoO assignment. For offspring with heterozygous genotype at a given variant site, the parental origin of the minor allele was determined using two methods; Mendelian and haplotype based methods as previously described (Figure 1C). The haplotype method was not performed when the number of polymorphic nucleotides observed in either parental or offspring haplotype was less than 5.

### Parent-of-origin association tests

Quantitative and binary traits were derived from electronic health records. Quantitative traits were normalized using rank inverse normal transformation prior to testing. The associations were tested using linear mixed models as implemented in BOLT^11^ for quantitative traits and SAIGE^13,15^ for binary traits, including covariates for age, age^2^, sex, age by sex interaction, indicator variable for genotyping array, and 10 principal components of ancestry calculated from array genotypes Traits were not included in downstream analyses or interpretation if there was no estimated heritability or if the genomic inflation factor was greater than 1.5 under additive or PoO models. To control multiplicity, variants were not tested if there were less than 5 heterozygous individuals of paternal inheritance or 5 heterozygous individuals of maternal inheritance with observed traits, assuming these variants have minimal power to reject the null hypothesis.

### Power simulations

The power estimates to detect PoO effect were simulated under the assumption that the true effects exhibit uniparental, polar dominance, or bipolar dominance patterns. The following parameters were used: total sample size = 134,049, offspring sample size = 22,051 (as in the DiscovEHR study), alpha = 5×10^−8^, and ranges of MAFs and effect sizes. The genotypic counts for a given MAF were derived assuming Hardy-Weinberg equilibrium. The phenotypes were simulated from a normal distribution with the assumed mean per genotype and standard deviation of 1. For the additive model, half of the heterozygous genotypic counts were simulated from paternal distribution while the other half were simulated from maternal distribution. The association between simulated genotypes and phenotypes was then tested under additive, parental, and differential models. The process was repeated 10,000 times and the proportion simulations yielding P-values more significant than alpha were used to estimate power.

## Data Availability

The data that support the reported findings may be available from the corresponding authors upon reasonable request.

## Acknowledgements

We thank the DiscovEHR participants and all research staff and teams at the Regeneron Genetics Center and Geisinger who contributed to the current study. We also thank the participants, researchers and organizers of the UK Biobank study. This work has been conducted using the UK Biobank application 26041. We thank the Regeneron postdoctoral program for their support and guidance for H.K.. The study is funded by Regeneron Genetics Center and Regeneron Pharmaceuticals.

## Author Contributions

H.K. conceived and conducted the study; B.Y. provided analytical support; C.V.H, A.R.S., J.S., C.G. contributed to the study design; J.S. contributed to the genetic relationship estimation. A.M. contributed to genotype imputation and provided computational support; C.V.H and A.R.S oversaw the study; H.K. wrote the manuscript; All authors reviewed the manuscript. Regeneron Genetics Center and Geisinger provided array genotype and phenotype data, respectively.

## Geisinger Regeneron DiscovEHR Collaboration Banner and Contribution Statement

Geisinger Banner and Contribution Statements

All authors/contributors are listed in alphabetical order.

Lance J. Adams^1^, Jackie Blank^1^, Dale Bodian^1^, Derek Boris^1^, Adam Buchanan^1^, David J. Carey^1^, Ryan

D. Colonie^1^, F. Daniel Davis^1^, Dustin N. Hartzel^1^, Melissa Kelly^1^, H. Lester Kirchner^1^, Joseph B. Leader^1^, David H. Ledbetter^1^, Ph.D., J. Neil Manus^1^, Christa L. Martin^1^, Michelle Meyer^1^, Tooraj Mirshahi^1^, Matthew Oetjens^1^, Thomas Nate Person^1^, Christopher Still^1^, Natasha Strande^1^, Amy Sturm^1^, Jen Wagner^1^, Marc Williams^1^

Contribution: Development and validation of clinical phenotypes used to identify study participants and (when applicable) controls.

Affiliations:

1. Geisinger, Danville, PA

Regeneron Genetics Center Banner and Contribution Statements

All contributors are listed in alphabetical order. RGC Management and Leadership Team:

Goncalo R. Abecasis, D.Phil.^1^, Aris Baras, M.D.^1^, Michael Cantor, M.D.^1^, Giovanni Coppola, M.D.^1^, Aris Economides, Ph.D.^1^, John D. Overton, Ph.D.^1^, Jeffrey G. Reid, Ph.D.^1^, Alan R. Shuldiner, M.D.^1^

Contribution: All authors contributed to securing funding, study design and oversight, and review and interpretation of data and results.

Sequencing and Lab Operations:

Christina Beechert^1^, Erin Brian^1^, Alex DeVito^1^, Caitlin Forsythe^1^, Erin D. Fuller^1^, Zhenhua Gu^1^, Joe LaRosa^1^, Michael Lattari^1^, Alexander Lopez^1^, Kia Manoochehri^1^, Justin Marcovici^1^, Manasi Pradhan^1^, John D. Overton, Ph.D.^1^, Thomas D. Schleicher^1^, Maria Sotiropoulos Padilla^1^, Karina Toledo^1^, Emelia Weihenig^1^, Louis Widom^1^, Sarah E. Wolf^1^, Ricardo H. Ulloa^1^

Contribution: Performed and are responsible for sample genotyping and exome sequencing, conceived and are responsible for laboratory automation, and responsible for sample tracking and the library information management system.

Genome Informatics:

Xiaodong Bai, Ph.D.^1^, Suganthi Balasubramanian, Ph.D.^1^, Leland Barnard, Ph.D.^1^, Andrew Blumenfeld^1^, Boris Boutkov^1^, Yating Chai, Ph.D.^1^, Gisu Eom^1^, Lukas Habegger, Ph.D.^1^, Young Hahn^1^, Alicia Hawes^1^, Shareef Khalid^1^, Olga Krasheninina^1^, Rouel Lanche^1^, Adam Mansfield^1^, Evan K. Maxwell, Ph.D.^1^, Mona Nafde^1^, Sean O’Keeffe, Ph.D.^1^, John Penn^1^, Ayesha Rasool^1^, William Salerno, Ph.D.^1^, Jeffrey C. Staples, Ph.D.^1^, Jeffrey G. Reid, Ph.D^1^

Contribution: Performed and are responsible for analysis needed to produce exome and genotype data, provided compute infrastructure development and operational support, provided variant and gene annotations and their functional interpretation of variants, and conceived and are responsible for creating, developing, and deploying analysis platforms and computational methods for analyzing genomic data.

Clinical Informatics:

Nilanjana Banerjee, Ph.D.^1^, Michael Cantor, M.D.^1^, Dadong Li Ph.D.^1^, Fabricio Sampaio Peres Kury M.D.^1^, Deepika Sharma B.H.M.S.^1^, Ashish Yadav^1^

Contribution: All authors contributed to the development and validation of clinical phenotypes used to identify study participants and (when applicable) controls.

Analytical Genomics and Data Science:

Goncalo R. Abecasis, D.Phil.^1^, Joshua Backman, Ph.D.^1^, Mathew Barber, Ph.D.^1^, Christian Benner, Ph.D.^1^, Shan Chen, Ph.D.^1^, Amy Damask, Ph.D.^1^, Manuel Allen Revez Ferreira, Ph.D.^1^, Lauren Gurski^1^, Jack Kosmicki, Ph.D.^1^, Alexander Li, Ph.D.^1^, Nan Lin, Ph.D.^1^, Daren Liu^1^, Jonathan Marchini Ph.D.^1^, Anthony Marcketta^1^, Joelle Mbatchou, Ph.D.^1^, Shane McCarthy, Ph.D.^1^, Colm O’Dushlaine, Ph.D.^1^, Charles Paulding, Ph.D.^1^, Claudia Schurmann, Ph.D.^1^, Dylan Sun^1^, Cristopher Van Hout, Ph.D.^1^, Kyoko Watanabe, Ph.D.^1^, Bin Ye^1^, Andrey Ziyatdinov, Ph.D.^1^

Contribution: Development of statistical analysis plans. QC of genotype and phenotype files and generation of analysis ready datasets. Development of statistical genetics pipelines and tools and use thereof in generation of the association results. QC, review and interpretation of result. Generation and formatting of results for manuscript figures.

Therapeutic Area Genetics:

Ariane Ayer^1^, Giovanni Coppola M.D.^1^, Silvio Alessandro Di Gioia, Ph.D.^1^, Jan Freudenberg, M.D.^1^, Sahar Gelfman, Ph.D.^1^, Claudia Gonzaga-Jauregui, Ph.D.^1^, Nehal Gosalia, Ph.D.^1^, Julie Horowitz, Ph.D.^1^, Luca Lotta M.D. Ph.D.^1^, Kavita Praveen, Ph.D.^1^

Contribution: Development of study design and analysis plans. Development and QC of phenotype definitions. QC, review, and interpretation of association results.

Functional Modeling:

Shek Man Chim, Ph.D.^1^, Giusy Della Gatta, Ph.D.^1^, Aris Economides, Ph.D.^3^, Lawrence Miloscio^1^, Harikiran Nistala, Ph.D.^1^, Trikaldarshi Persaud^1^

Contribution: Development of *in vivo* and *in vitro* experimental biology and interpretation. Planning, Strategy, and Operations:

Paloma M. Guzzardo, Ph.D.^2^, Marcus B. Jones, Ph.D.^2^, Michelle LeBlanc, Ph.D.^2^, Jason Mighty, Ph.D.^2^, Lyndon J. Mitnaul, Ph.D.^2^

Contribution: Contributed to the management and coordination of all research activities, planning and execution, managed the review of the project.

Affiliations:

^1^ Regeneron Genetics Center, Tarrytown, NY USA, ^2^ Regeneron Pharmaceuticals, Tarrytown, NY USA

## References

1. Lawson, H.A., Cheverud, J.M. & Wolf, J.B. Genomic imprinting and parent-of-origin effects on complex traits. Nat Rev Genet 14, 609–17 (2013).

2. Kong, A. et al. Parental origin of sequence variants associated with complex diseases. Nature 462, 868–74 (2009).

3. Wallace, C. et al. The imprinted DLK1-MEG3 gene region on chromosome 14q32.2 alters susceptibility to type 1 diabetes. Nat Genet 42, 68–71 (2010).

4. Small, K.S. et al. Identification of an imprinted master trans regulator at the KLF14 locus related to multiple metabolic phenotypes. Nat Genet 43, 561–4 (2011).

5. Hanson, R.L. et al. Strong parent-of-origin effects in the association of KCNQ1 variants with type 2 diabetes in American Indians. Diabetes 62, 2984–91 (2013).

6. Perry, J.R. et al. Parent-of-origin-specific allelic associations among 106 genomic loci for age at menarche. Nature 514, 92–97 (2014).

7. Zoledziewska, M. et al. Height-reducing variants and selection for short stature in Sardinia. Nat Genet 47, 1352–1356 (2015).

8. Benonisdottir, S. et al. Epigenetic and genetic components of height regulation. Nat Commun 7, 13490 (2016).

9. Connolly, S., Anney, R., Gallagher, L. & Heron, E.A. A genome-wide investigation into parent-of-origin effects in autism spectrum disorder identifies previously associated genes including SHANK3. Eur J Hum Genet 25, 234–239 (2017).

10. Mozaffari, S.V. et al. Parent-of-origin effects on quantitative phenotypes in a large Hutterite pedigree. Commun Biol 2, 28 (2019).

11. Zink, F. et al. Insights into imprinting from parent-of-origin phased methylomes and transcriptomes. Nat Genet 50, 1542–1552 (2018).

12. Morison, I.M., Ramsay, J.P. & Spencer, H.G. A census of mammalian imprinting. Trends Genet 21, 457–65 (2005).

13. Loh, P.R. et al. Efficient Bayesian mixed-model analysis increases association power in large cohorts. Nat Genet 47, 284–90 (2015).

14. Bastepe, M. The GNAS locus and pseudohypoparathyroidism. Adv Exp Med Biol 626, 27–40 (2008).

15. Zhou, W. et al. Efficiently controlling for case-control imbalance and sample relatedness in large-scale genetic association studies. Nat Genet 50, 1335–1341 (2018).

16. Monk, D., Mackay, D.J.G., Eggermann, T., Maher, E.R. & Riccio, A. Genomic imprinting disorders: lessons on how genome, epigenome and environment interact. Nat Rev Genet 20, 235–248 (2019).

17. Zeng, Y. et al. Parent of origin genetic effects on methylation in humans are common and influence complex trait variation. Nat Commun 10, 1383 (2019).

18. Wolf, J.B. & Wade, M.J. What are maternal effects (and what are they not)? Philos Trans R Soc Lond B Biol Sci 364, 1107–15 (2009).

19. Gleason, G. et al. The serotonin1A receptor gene as a genetic and prenatal maternal environmental factor in anxiety. Proc Natl Acad Sci U S A 107, 7592–7 (2010).

20. Kong, A. et al. The nature of nurture: Effects of parental genotypes. Science 359, 424–428 (2018).

21. Dewey, F.E. et al. Distribution and clinical impact of functional variants in 50,726 whole-exome sequences from the DiscovEHR study. Science 354(2016).

22. McCarthy, S. et al. A reference panel of 64,976 haplotypes for genotype imputation. Nat Genet 48, 1279–83 (2016).

23. Bycroft, C. et al. The UK Biobank resource with deep phenotyping and genomic data. Nature 562, 203–209 (2018).

24. Purcell, S. et al. PLINK: a toolset for whole-genome association and population-based linkage analysis. Am J Hum Genet 81, 559–575 (2007).

25. Staples, J. et al. PRIMUS: rapid reconstruction of pedigrees from genome-wide estimates of identity by descent. Am J Hum Genet 95, 553–564 (2014).

